# Multi-trait Analysis of GWAS (MTAG) of Substance Use Traits Identifies Novel Genetic Loci and Phenomic Associations

**DOI:** 10.1101/2022.07.06.22277340

**Authors:** Heng Xu, Sylvanus Toikumo, Richard C. Crist, Klaudia Glogowska, Joseph D. Deak, Joel Gelernter, Emma C. Johnson, Henry R. Kranzler, Rachel L. Kember

## Abstract

**Introduction:** A large majority of genome-wide significant (GWS) loci identified for substance use traits (SUTs) in genome-wide association studies (GWAS) have been for alcohol and smoking-related phenotypes. GWAS of opioid use disorder (OUD) and cannabis use disorder (CUD) have lagged those of the two historically legal substances.

**Methods:** We applied multi-trait analysis of GWAS (MTAG) to 2,888,727 single nucleotide polymorphisms (SNPs) common to GWAS of four SUTs (OUD, CUD, alcohol use disorder [AUD], and smoking initiation [SMK]) in European-ancestry (EUR) subjects. We calculated polygenic risk scores (PRS) for the four traits in an independent sample (i.e., the Yale-Penn sample; N=5,692 EUR) and examined the power increment for each set of MTAG-GWAS summary statistics relative to those of the input GWAS.

**Results:** MTAG increased the effective sample size for all four SUTs, which showed high pairwise genetic correlations. After clumping, MTAG identified independent GWS SNPs for all 4 traits: 41 SNPs in 36 loci (including 5 novel loci not previously associated with any SUT) for OUD; 74 SNPs in 60 loci (including 4 novel loci) for CUD; 63 SNPs in 52 loci (including 10 novel loci) for AUD; and 183 SNPs in 144 loci (including 8 novel loci) for SMK. In PRS analyses in the Yale-Penn sample, the MTAG-derived PRS consistently yielded more significant associations with both the corresponding substance use disorder diagnosis and multiple related phenotypes than each of the 4 GWAS-derived PRS.

**Conclusions:** MTAG boosted the number of GWS loci for the 4 SUTs, including identifying genes not previously linked to any SUT. MTAG-derived PRS also showed stronger associations with expected phenotypes than PRS for the input GWAS. MTAG can be used to identify novel associations for SUTs, especially those with sample sizes smaller than for historically legal substances.

## Introduction

Genome-wide association studies (GWAS) of substance use traits (SUTs) have successfully identified genome-wide significant (GWS) risk variants. However, despite high estimates of heritability (∼50%) for many SUTs, difficulty in recruiting large study samples has limited gene discovery efforts ^1^. Because common genetic variants have small effects on trait susceptibility, large samples are usually needed to provide adequate statistical power to identify GWS variant associations. Thus, variant discovery for some traits [e.g., opioid use disorder (OUD) and cannabis use disorder (CUD)] has lagged that of more common SUTs, including alcohol and smoking-related phenotypes.

Recent GWAS of OUD, CUD, alcohol use disorder (AUD), and smoking initiation (SMK) have identified more than 300 substance-associated loci, some of which are shared between multiple SUTs. SUTs have also shown high degrees of genetic correlation in both GWAS and twin and family studies of the respective traits ^2^. A GWAS of OUD in the Million Veteran Program (MVP), with 31,473 OUD cases, identified 10 loci in the cross-ancestry meta-analysis ^3^ including *OPRM1, FURIN, NCAM1*, and 7 other novel genes. An initial GWAS of cannabis use disorder (CUD) identified one GWS locus (on chromosome 8, near *CHRNA2* and *EPHX2*) ^4^, which was subsequently confirmed in a meta-analysis that included 20,916 CUD cases and identified a second GWS locus on chromosome 7 (*FOXP2*) ^5^. A cross-ancestry GWAS of AUD in the MVP sample found 26 associated loci, of which 4 (*ZNF804A, MLN, NICN1, MIR5694*), though previously associated with psychiatric or other SUTs, were novel for alcohol-related phenotypes ^6^. Finally, a meta-analysis of smoking initiation (SMK) in 1.2 million individuals identified 278 loci (including *NCAM1*) ^7^.

Multi-trait analysis of GWAS (MTAG), developed to boost the statistical power of GWAS by incorporating information from effect estimates across traits ^8^, enables the joint analysis of multiple, genetically correlated traits. MTAG is also valuable because it can be applied to summary statistics of GWAS rather than requiring individual genotypes, addressing the sample overlap often present across GWAS discovery samples for different traits using linkage disequilibrium (LD) score regression ^8^. Further, it generates trait-specific estimates of the effects of each single nucleotide polymorphism (SNP), yielding a lower mean-squared error than standard, single-trait GWAS estimates when a key assumption of the procedure—that all SNPs share a variance-covariance matrix of effect sizes across traits—is not satisfied ^8^.

MTAG has been employed to boost genetic findings for SUTs, both for traits involving the same substance and for cross-substance traits. In a meta-analysis of problematic alcohol use (PAU) ^9^, MTAG analysis of PAU and a measure of weekly alcohol consumption ^7^ increased the number of independent loci for PAU from 29 to 76. MTAG analysis of OUD with AUD and CUD ^10^ increased the number of GWS loci for OUD to 18 from 3 in the initial GWAS meta-analysis. Because MTAG for multiple, genetically correlated SUTs yields greater statistical and interpretive power than individual trait GWAS, the availability of large GWAS of alcohol- and smoking-related traits could enhance findings from GWAS of traits with smaller accumulated samples (e.g., CUD and OUD).

Here we conduct an MTAG analysis of the largest available GWAS for four SUTs: OUD, CUD, AUD, and SMK. We integrated information from the four sets of GWAS summary statistics to identify novel loci for each SUT. We also conducted gene prioritization, gene-set, and protein-protein interaction analyses to characterize the underlying biology of the novel genes in the context of SUTs. Finally, we generated polygenic risk scores (PRS) to examine the power increment for each set of MTAG-GWAS summary statistics in an independent sample.

## Methods

### GWAS Summary Statistics

We examine four substance use traits (SUTs) in European ancestry (EUR) samples: OUD ^3^ (Effective N = 74,635), CUD ^5^ (Effective N = 48,900), AUD ^6^ (Effective N = 171,601), and SMK ^7^ (Effective N = 632,802). Both the OUD and AUD GWAS were conducted in the Million Veteran Program (MVP), a large genomic dataset linked to electronic health records data. The CUD GWAS was a meta-analysis of three datasets (Psychiatric Genomics Consortium, iPSYCH, deCODE). The SMK GWAS was a meta-analysis of multiple datasets conducted by the GWAS consortium of alcohol and nicotine use (GSCAN). Because our downstream PRS analysis used the Yale-Penn dataset, and the CUD GWAS included genotype data from Yale-Penn subjects, we avoided sample overlap by generating summary statistics for CUD using a “leave-one-out” meta-analysis that excluded Yale-Penn subjects. All other GWAS were independent of the Yale-Penn dataset.

#### Genetic Correlations

We calculated pairwise genetic correlations (*r*_*g*_) for the four SUTs using linkage disequilibrium score regression (LDSC) ^11^ and HapMap 3 SNPs. Pre-computed European LD scores and weights were downloaded from the LDSC GitHub website (https://github.com/bulik/ldsc).

### Multi-trait Analysis of GWAS Summary Statistics (MTAG)

Given the high pairwise genetic correlations for the four SUTs, which ranged from 0.45 to 0.80, we conducted a joint analysis of these traits using MTAG, a generalized meta-analysis method that outputs trait-specific SNP associations. It uses bivariate LD score regression to account for potential sample overlap between two or more input summary statistics. We used trait-specific effective sample sizes and transformed Z-scores. SNPs present in all four sets of SUT summary statistics were included in the MTAG calculation. Default MTAG parameters were used (i.e., each SNP’s sample size must be larger than two-thirds of the 90^th^ percentile of all SNPs’ sample sizes). We used PLINK1.9 to perform clumping procedures on the four MTAG-GWAS results across a range of 3000 kb and r^2^ > 0.1. GWS variants located within 1 Mb were merged into a single locus. Loci were annotated with the nearest protein-coding gene (within 1 Mb) using SNPsnap ^12^. We calculated maxFDR ^8^ as the upper bound of FDR for each MTAG-GWAS result. Low maxFDR values support the robustness of the MTAG-GWAS results. Of note, SNP rs1229984, located in the alcohol dehydrogenase gene *ADH1B*, did not pass the default MTAG quality control parameter as it was not present in at least two-thirds of the sample. Because of the well-known strong association of rs1229984 with alcohol phenotypes [6,9], we conducted a separate analysis with a less stringent filter to include that SNP and report the results in Supplemental Table 18.

### Identification of novel variants for each SUT

We systematically evaluated whether variants identified in the MTAG analysis were previously associated with either the primary SUT or other SUTs. First, for each lead variant, we determined whether any nearby variant (within 1 Mb) was GWS in the initial GWAS or in the other three contributing GWAS. We then determined whether the locus had prior SUT associations using the GWAS catalog ^13^ implemented in FUMA ^14^, annotating all lead variants with prior associations with any SUT (not limited to the GWAS included in this study). For completeness, we also labelled the variant with the closest protein-coding gene (within 1 Mb) and searched the GWAS catalog ^13^ for prior associations of that gene with SUTs. Loci that were not previously GWS for any SUT were labeled as “novel”.

### Gene-Set Analysis

To determine whether the identified genes are involved in important biological processes, GWS SNPs were mapped to genes by ANNOVAR ^15^ implemented in FUMA ^14^. Next, we curated gene-set enrichment and Gene Ontology (GO) terms using the GO annotation database ^16,17^, with gene-set enrichment p-values adjusted using a Bonferroni correction for each test.

### Protein-Protein Interaction

We used STRING database v11.5 ^18^ to conduct protein-protein interaction (PPI) analyses. For each SUT MTAG result, we used annotated GWS genes as input to query the PPIs in the database. PPI enrichment p-value and pairwise interaction scores were reported by the STRING database. We used a cut-off interaction score >0.4 to identify PPIs with high confidence.

### Yale-Penn Dataset

The Yale-Penn sample was recruited for genetic studies of substance use disorders (SUDs) ^19^. It was deeply phenotyped using the Semi-Structured Assessment for Drug Dependence and Alcoholism (SSADDA), a comprehensive psychiatric interview schedule that assesses the physical, psychosocial, and psychiatric manifestations of SUDs and co-occurring psychiatric disorders ^20,21^. Using the SSADDA, trained interviewers elicit information on demographics, substance use history, psychosocial history, medical history, and lifetime diagnostic criteria for DSM-IV ^22^ and DSM-5 ^23^ substance use and DSM-IV psychiatric disorders. Diagnoses and individual criteria for SUDs and psychiatric disorders obtained with the SSADDA show acceptable reliability ^20,21^. The Yale-Penn PheWAS dataset includes 5,692 unrelated European-ancestry genotyped individuals and over 650 summarized phenotypes categorized into 20 substance, medical, demographic, and psychiatric sections ^19^.

### Polygenic Risk Score (PRS) and Phenotype Association Test

We used PRS-continuous shrinkage (PRS-CS) ^24^ to generate PRS in the Yale-Penn dataset ^19^, pre-computed LD reference for HapMap3 SNPs in EUR samples to account for local LD, and an optimal global shrinkage parameter learned from the data. We generated four GWAS-based PRS and four MTAG-based PRS, i.e., one of each for each of the four SUTs. To ensure comparability between GWAS-based and MTAG-based PRS, only the SNPs used in the MTAG calculation were included in PRS calculation. Effective sample size was used to generate GWAS- and MTAG-based PRS.

To compare the power of GWAS-based PRS with MTAG-based PRS, we tested the associations of each PRS in the Yale-Penn dataset with phenotypes in the corresponding substance section (e.g., AUD PRS were tested with alcohol-related phenotypes). We used linear regression or logistic regression models as appropriate, covarying for age, sex, and 10 genetic principal components. Each substance section includes DSM-IV and DSM-5 diagnoses for the corresponding substance use disorder. For diagnoses, we calculated the incremental pseudo *R*^2^ value after adding the polygenic score to the logistic regression model.

## Results

### Genetic Correlation Between SUTs

Pairwise *r*_*g*_ calculated using LDSC were significant and were moderate or high across all pairs of SUTs (Figure 1, Supplementary Table 1). The strongest *r*_*g*_ was between OUD and AUD (0.80), followed by OUD and CUD (0.67), CUD and SMK (0.65), CUD and AUD (0.63), AUD and SMK (0.52) and OUD and SMK (0.45).

**Figure 1:**
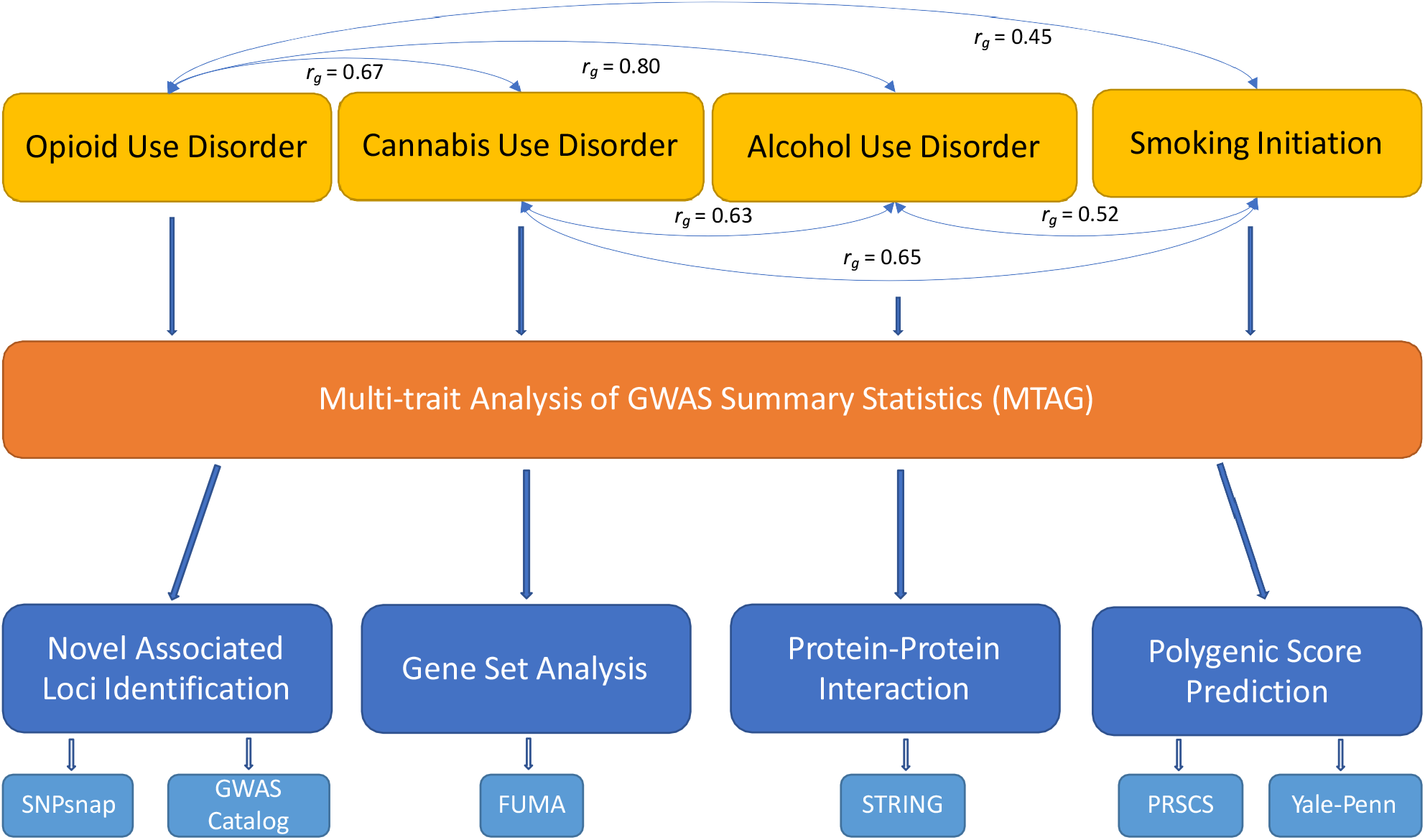
Overview of the analysis. The four SUTs that were used in MTAG-analysis have pair-wise genetic-correlations between 0.45 to 0.80. For each SUT MTAG-GWAS result, we identified novel loci, performed gene-set analysis, protein-protein interaction analysis, and examined the increased predictive power of the corresponding polygenic risk score.

### MTAG SUTs and Locus Discovery

We retained 2,888,727 SNPs for the MTAG analysis after extracting common SNPs from all four input sets of GWAS summary statistics and filtering them with the criteria described in the MTAG methods description. The mean χ^2^ statistics for the MTAG-GWAS results are: χ^2^ _*MTAG-OUD*_ = 1.38, χ^2^ _*MTAG-CUD*_ = 1.46, χ^2^ _*MTAG-AUD*_ = 1.46 and χ^2^ _*MTAG-SMK*_ = 1.87. The maxFDR values are 6.7 × 10^−2^ for MTAG-OUD, 1.1 × 10^−1^ for MTAG-CUD, 8.9 × 10^−3^ for MTAG-AUD and 1.2 × 10^−4^ for MTAG-SMK (Table 1).

**Table 1.**
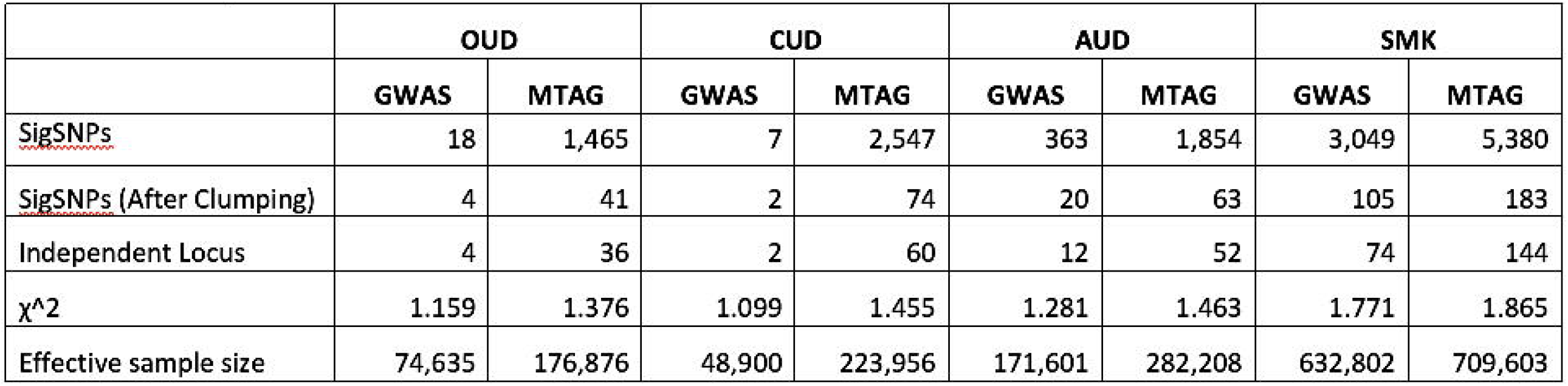
Summary of MTAG Results

Among the four sets of MTAG results (Figure 2, Supplementary Tables 2-5), 20 loci were significantly associated with all four SUTs, the most significant of which was the locus containing *NCAM1*, supported by two intronic variants in complete LD (rs1940701 for OUD and AUD and rs4479020 for CUD and SMK). Other loci associated with all four SUTs included intronic variants in *DPP4* (rs6432708) and *CADM2* (rs62250713) and an intergenic variant near *ZNF184* (rs35984974).

**Figure 2:**
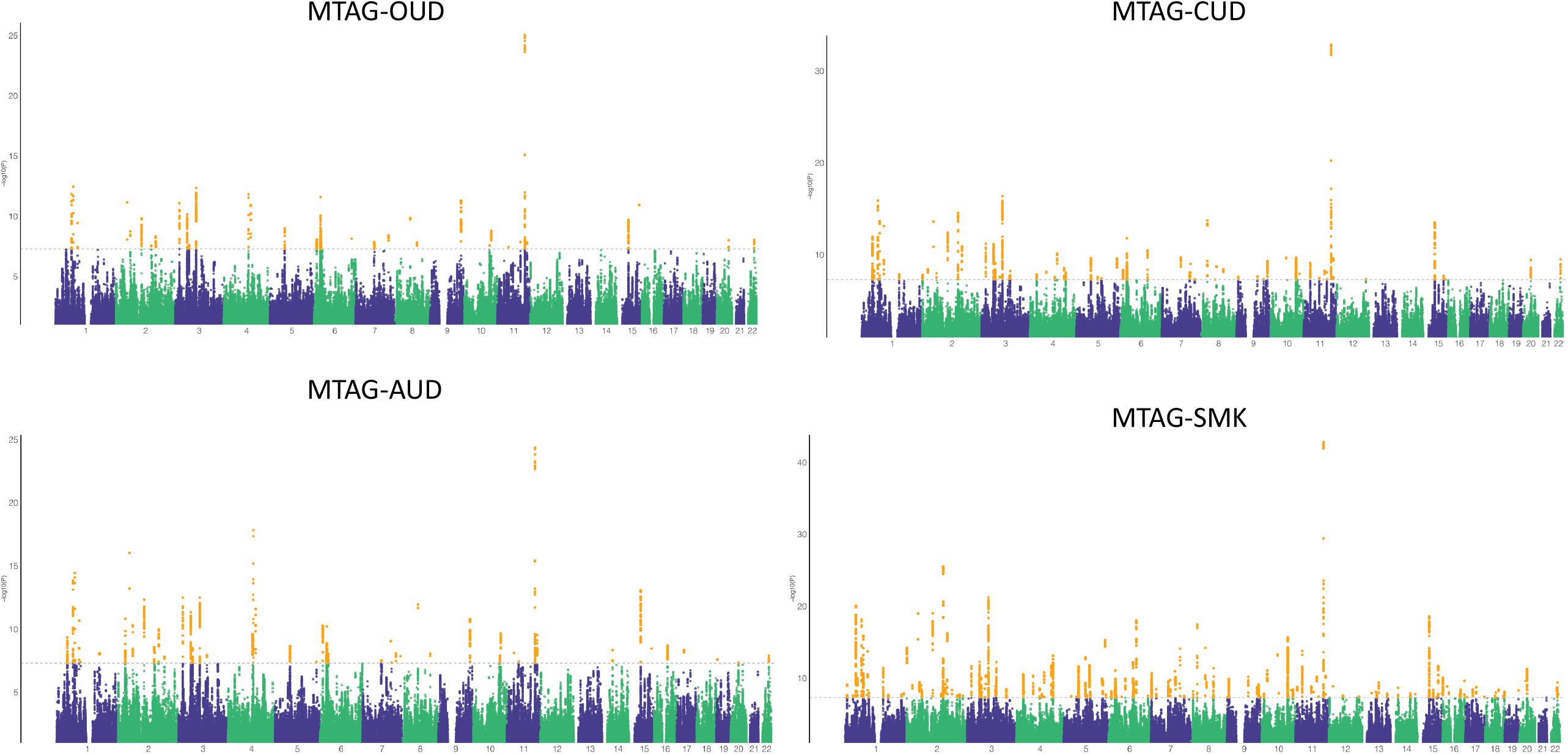
Manhattan plots of MTAG-OUD (N_Effective_ = 176,876), MTAG-CUD (N_Effective_ = 223,956), MTAG-AUD (N_Effective_ = 282,208) and MTAG-SMK (N_Effective_ = 709,603). Dashed-lines indicate genome-wide significance (P < 5 × 10^−8^) and yellow dots indicate genome-wide significance SNPs.

### Opioid use disorder (MTAG-OUD)

MTAG increased the effective sample size for OUD from 74,635 in the input GWAS to 176,876 in the MTAG analysis. After clumping, we identified 41 independent GWS SNPs in 36 genomic risk loci (Supplementary Table 2). Of these, 3 loci were GWS in the OUD GWAS, 17 were GWS in at least one of the other of the three SUT GWAS used for MTAG, and 30 were GWS in other GWAS of SUTs, not limited to the GWAS included in this study. Variants including exonic SNPs in *OPRM1* (rs1799971, p = 7.52 × 10^−9^) and *FURIN* (rs4702, p = 3.29 × 10^−12^) have been shown to be GWS in previous OUD GWAS ^3,10^. Five GWS loci are novel with no prior associations found with OUD or any SUT. These include a variant in the 3’ UTR of *POR* (rs17685, p = 1.42 × 10^−8^), intronic variants in *CNOT4* (rs2696880, p = 3.97 × 10^−9^) and *MTMR2* (rs7110786, p = 1.38 × 10^−8^), and intergenic variants near *TMEM170B* (rs112126124, p = 8.35 × 10^−9^) and *SNAI1* (rs73274724, p = 9.92 × 10^−9^).

### Cannabis use disorder (MTAG-CUD)

MTAG increased the effective sample size from 48,900 to 223,956, yielding 74 independent GWS SNPs in 60 genomic risk loci (Supplementary Table 3). Of the 60 loci, 3 were GWS in the CUD GWAS, 43 were GWS in at least one of the other three SUT GWAS used for MTAG, and 51 were GWS in other GWAS of SUTs. We replicated previously associated loci for CUD ^5^, including an intergenic variant near *CLU* in the same locus identified in the input CUD GWAS, an intronic variant in *GBF1* (rs4919626, p = 1.38 × 10^−8^), and an exonic variant in *FURIN* (rs4702, p = 4.69 × 10^−8^). Four novel GWS loci with no prior associations with any SUT were identified, including three intronic variants - one each in *CNOT4* (rs2696880, p = 4.44 × 10^−8^), *TMEM245* (rs11794648, p = 4.26 × 10^−8^), and *MTMR2* (rs7110786, p = 8.21 × 10^−9^) - and an intergenic variant near *TMEM170B* (rs112126124, p = 4.47 × 10^−9^).

### Alcohol Use Disorder (MTAG-AUD)

With an effective sample size that increased from 171,601 in the input GWAS to 282,208 in the MTAG analysis, we identified 63 independent GWS SNPs in 52 genomic risk loci for AUD (Supplementary Table 4). Of these, 9 loci were GWS in the AUD GWAS, 23 were GWS in at least one of the other three SUT GWAS used for MTAG, and 40 were GWS in other GWAS of SUTs. These included multiple previously reported AUD-associated variants – exonic variants in *GCKR* (rs1260326, p = 1.53 × 10^−11^) and *SLC39A8* (rs13107325, p = 1.47 × 10^−18^), and intronic variants in *ANKK1* (rs12360992, p = 3.29 × 10^−8^) and *FTO* (rs7206122, p = 2.03 × 10^−9^) ^6^. We also identified 10 novel GWS loci, including exonic variants in *POR* (rs17685, p = 3.44 x10^−8^) and *SYNGAP1* (rs411136, p = 9.10 × 10^−9^); intronic variants in *DNM3* (rs742510, p = 9.63 × 10^−9^), *CSMD3* (rs6469450, p = 8.91 × 10^−9^) *CNOT4* (rs2686880, p = 8.26 × 10^−9^), *LRFN5* (rs58734839, p = 4.55 × 10^−9^), *ZNF804A* (rs1366839, p = 1.53 × 10^−8^), and *TCF20* (rs9306356, p = 1.30 × 10^−8^); and two intergenic variants near *TMEM170B* (rs112126124, p = 5.62 × 10^−11^) and *SORCS3* (rs73274724, p = 1.27 × 10^−8^).

### Smoking Initiation (MTAG-SMK)

With an effective sample size increase from 637,082 to 709,603, MTAG identified 183 independent GWS SNPs in 144 genomic risk loci (Supplementary Table 5). Of these, 86 were GWS in the SMK GWAS, 7 were GWS in at least one of the other three SUT GWAS used for MTAG, and 130 were GWS in other GWAS of SUTs. Eight GWS loci were novel, including a variant within *TNRC6B* (rs5750911, p = 2.38 × 10^−9^) and seven intergenic variants: one each near *WDR12* (rs4675308, p = 2.50 × 10^−8^), *PCDH7* (rs7680926, p = 8.81 × 10^−9^), *ITGA1* (rs7680926, p = 4.05 × 10^−8^), *TMEM170B* (rs12526369, p = 7.41 × 10^−9^), *SP4* (rs6974377, p = 3.37 × 10^−9^), *CTDP1* (rs56235016, p = 7.76 × 10^−9^), and *SORCS3* (rs10884186, p = 4.01 × 10^−8^).

### Gene-Set Analysis

After Bonferroni correction, we identified enriched gene sets for all studied traits including 28 for MTAG-OUD, 73 for MTAG-CUD, 51 for MTAG-AUD, and 70 for MTAG-SMK, as shown in Supplementary Tables 6-9. The gene-set “regulation of cell differentiation”, which contains the novel gene *POR* and the opioid-specific gene *OPRM1*, showed significant enrichment for OUD (P_Bon_ = 1.0 × 10^−3^). “Protein dimerization activity”, one of the significantly enriched gene-sets for CUD (P_Bon_ = 3.7 × 10^−9^), harbors the novel genes *MTMR2* and *FOXP2*. For AUD, the enriched gene set “regulation of synapse structure or activity” (P_Bon_ = 1.2 × 10^−2^) contained the novel genes *DNM3, LRFN5*, and *SYNGAP1* along with *DRD2*. For SMK, 70 gene sets showed significant enrichment, with the novel gene *TNRC6B* mapping to 12 of them. Of note, the gene set “neuron differentiation”, which includes *NCAM1*, is significantly enriched (P_Bon_ = 3.1x 10^−2^).

### Protein-protein Interaction

Using the STRING database, we observed significant PPI enrichment (p < 0.05) for genes that were GWS for MTAG-AUD and MTAG-SMK, whereas PPI enrichment was non-significant for GWS genes in MTAG-OUD (p = 0.175) and MTAG-CUD (p = 0.175) (Supplementary Tables 10-13). For GWS genes in MTAG-AUD, we identified high PPI for *NF1* and *SYNGAP1* (interaction score = 0.697), *NF1* and *CSMD3* (interaction score = 0.42), and *NCAM1* and *SEMA6D* (interaction score = 0.467). For GWS genes in MTAG-SMK, *TNRC6B* and *FOXO3*, two smoking-associated genes, showed high PPI (interaction score = 0.9).

### PRS Associations with Primary Phenotypes

As shown in Figure 3, all four MTAG-based PRS (PRS_MTAG_) showed stronger associations with the primary diagnosis than the individual-trait GWAS-based PRS (PRS_GWAS_) (Supplementary Tables 14-17). This difference was most evident for DSM-IV cannabis dependence, where the Bonferroni-corrected association was non-significant with PRS_GWAS-CUD_ (OR = 1.16, p = 1.94 x10^−3^) but significant with PRS_MTAG-CUD_ (OR = 1.30, p = 2.18 × 10^−9^).

**Figure 3:**
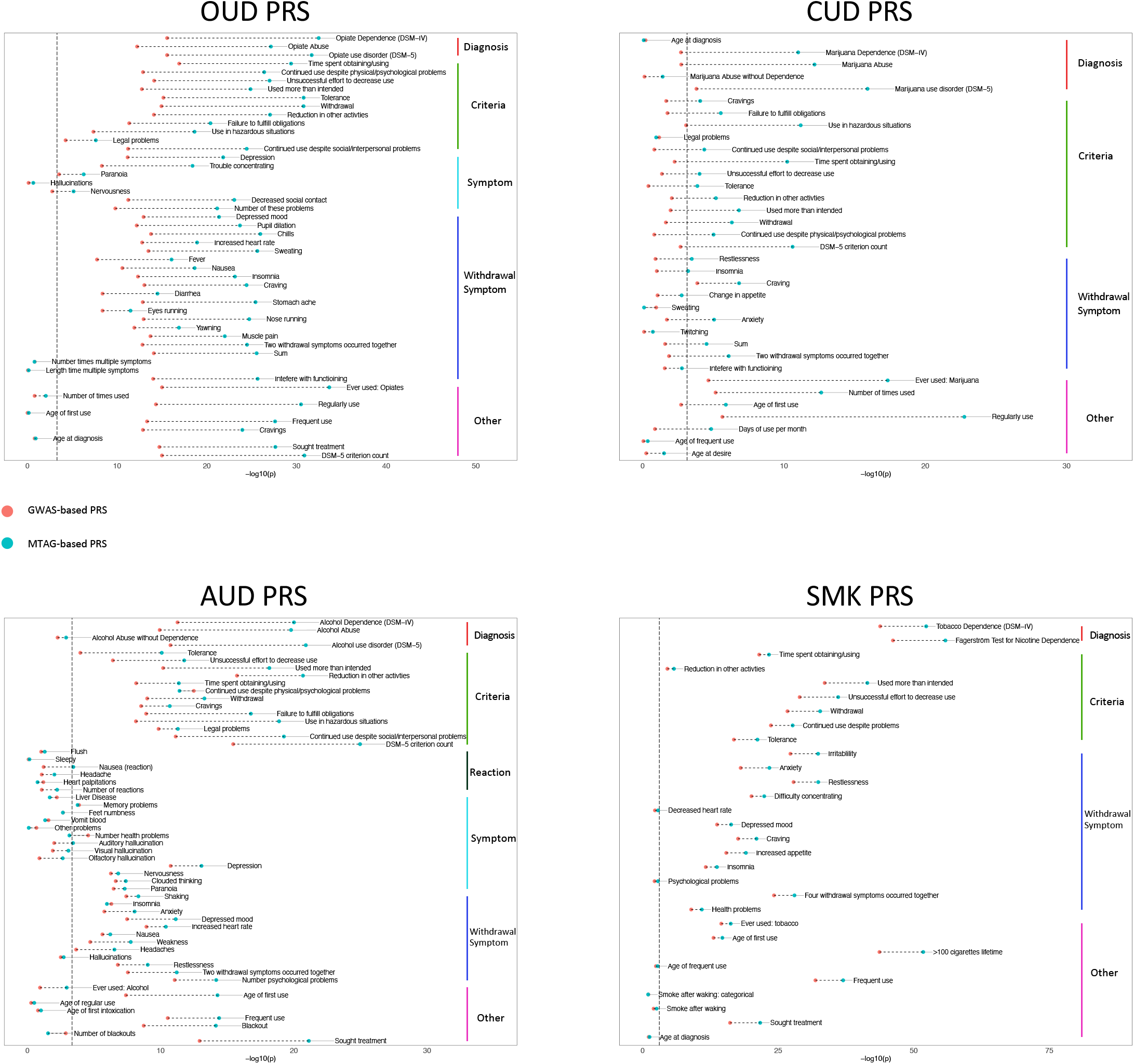
Comparison of GWAS-based PRS and MTAG-based PRS. Red and blue dots in each plot represent the GWAS-based polygenic risk score and MTAG-based polygenic risk score respectively. Vertical dashed lines indicate significance threshold after Bonferroni correction.

Associations with the corresponding primary diagnoses for the other three SUTs were all orders of magnitude less significant for the PRS_GWAS_ than the PRS_MTAG_: PRS_GWAS-OUD_: OR = 1.30, p = 2.64 x10^−16^ vs. PRS_MTAG-OUD_: OR = 1.50, p = 3.34 x10^−33^; PRS_GWAS-AUD_: OR = 1.27, p = 5.44 x10^−12^ vs. PRS_MTAG-AUD_: OR = 1.40, p = 9.94 x10^−21^; PRS_GWAS-SMK_: OR = 1.61, p = 1.57 x10^−44^ vs. PRS_MTAG-SMK_: OR = 1.71, p = 5.14 x10^−53^.

For each of the SUTs, there were more Bonferroni-corrected significant associations with PRS_MTAG_ than PRS_GWAS_ (Supplementary Tables 14-17). Notably, the number of significantly associated phenotypes for PRS_GWAS-CUD_ was six, while for PRS_MTAG-CUD_ it was 25. Phenotypes that became significantly associated when using PRS_MTAG-CUD_ included age of first use of marijuana, the DSM-IV cannabis dependence diagnosis, and the DSM-5 CUD criterion count.

In addition, the incremental *R*^*2*^ values both for diagnoses and related phenotypes were higher for PRS_MTAG_ than PRS_GWAS_ (Table 2). The greatest improvement was between PRS_MTAG-CUD_ and PRS_GWAS-CUD_, where the incremental *R*^*2*^ for “ever used cannabis” was 1.62% and 0.38%, respectively. Moderate improvement was also observed for PRS_MTAG-SMK_ compared with the well-powered PRS_GWAS-SMK_.

**Table 2.** PRS Increment R^2^

|  |  | GWAS-PRS | MTAG-PRS |
| --- | --- | --- | --- |
| Opioid | Ever Used Opioid | 0.97% | 2.31% |
|  | DSM-IV Diagnosis | 1.03% | 2.26% |
|  | DSM-5 Diagnosis | 1.01% | 2.16% |
| Cannabis | Ever Used Cannabis | 0.38% | 1.62% |
|  | DSM-IV Diagnosis | 0.19% | 0.91% |
|  | DSM-5 Diagnosis | 0.23% | 1.13% |
| Alcohol | Ever Used Alcohol | 0.26% | 1.11% |
|  | DSM-IV Diagnosis | 0.91% | 1.68% |
|  | DSM-5 Diagnosis | 0.84% | 1.72% |
| Smoking | Ever Used Smoking | 1.81% | 2.04% |
|  | DSM-IV Diagnosis | 3.43% | 4.16% |

## Discussion

We performed a joint analysis of four SUTs using MTAG, which yielded an effective sample size that was up to four-fold that of the four individual GWAS. This led to our identifying novel associated variants in loci not previously linked to any SUT. Among the most significant risk loci for substance-use phenotypes are those that encode proteins with clear connections to the substance involved. This includes the mu-opioid receptor for OUD ^25^, various nicotinic cholinergic receptor subunits for smoking initiation ^7^, and alcohol metabolism enzymes for alcohol consumption and AUD ^26^.

However, beyond the substance-specific proteins that directly interact with the drug are a wide range of biological mechanisms common to all addictive behavior, which includes reward pathways, learning and memory, withdrawal, and other functions^27^. We therefore expected that the greater statistical power of MTAG would reveal novel genes with associations to multiple substances based on common mechanisms of risk (i.e., an addiction factor ^28,29^). In fact, five of the 19 novel loci identified in our analysis were significantly associated with two or more substances. Variants in *POR* were significant for OUD and AUD, in *MTMR2* was significant for OUD and CUD, near *SORCS3* was significant for AUD and SMK, in *CNOT4* was significant for OUD, CUD and AUD, and near *TMEM170B* was significant for all four SUTs. Interestingly, *CNOT4* was recently identified in a GWAS of maximum alcohol use ^30^, which supports our discovery here. The use of multiple substances is common and has been associated with poorer treatment outcomes in individuals with SUDs ^31^. These shared genes may represent targets for therapies aimed at treating co-occurring SUDs.

*POR* encodes a reductase that contributes electrons to cytochrome P450 enzymes ^32^, which are essential components of drug metabolism. Variation in *POR* can affect the enzymatic activities of multiple CYP450 enzyme family members ^33–35^, including CYP2D6 and CYP3A4 ^33^, enzymes responsible for the primary metabolism of many opioids. Our MTAG results indicate that the A-allele of the *POR* variant rs17685 is associated with decreased risk of OUD and AUD. This SNP was also found to be an expression quantitative trait locus (eQTL) for *POR*, with the A-allele predicting higher expression in cerebellum ^36^. If increased POR correlates with increased CYP450 activity, the association of rs17685 with OUD could reflect altered opioid pharmacokinetics. There may be a similar connection between *POR* and AUD. CYP2E1, which accounts for ∼20% of alcohol metabolism ^37,38^, has been shown to interact with POR ^39^. CYP2E1 expression is also induced by ethanol itself ^38^. Thus, there is a potential synergistic effect wherein rs17685-A allele carriers who consume alcohol may simultaneously have an increased amount and activity of the enzyme.

We also identified novel loci associated with individual SUTs. For OUD, we detected a novel variant near *SNAI1* and for CUD, a novel intronic variant in *TMEM245*. For AUD, six variants were identified that had not previously been associated with any SUT (*SYNGAP1, DNM3, CSMD3, LRFN5, ZNF804A, TCF20*). For SMK, six novel variants not associated with any other SUTs were identified (*TNRC6B, WDR12, PCDH7, ITGA1, SP4, CTDP1*).

In addition to polysubstance use, patients with SUDs also have higher rates of comorbid psychiatric disorders than the general population ^40^. Phenome-wide association studies (PheWAS) using data from electronic health records (EHR) have shown associations between PRS for SUTs and non-substance use psychiatric diagnostic codes ^9^, suggesting genetic overlap between them. Consistent with this hypothesis, nine of the 19 novel genes identified in our MTAG analysis were significant in GWAS of depression or schizophrenia ^41–45^.

A prior MTAG analysis of PAU^9^, which leveraged information from a GWAS of drinks per week, identified 119 GWS variants. Of the variants in common between that MTAG analysis and the present analysis, we also observed association with 14 of the 45 SNPs in our MTAG-AUD results. In our MTAG-OUD analysis, we also replicated six of the nine variants in common with a recent OUD MTAG analysis of OUD, CUD and AUD ^10^. Although *OPRM1* was not significantly associated in the prior MTAG, our MTAG-OUD analyses identified the *OPRM1* SNP rs1799971 as a lead variant, potentially due to our inclusion of the smoking initiation GWAS or the larger input OUD GWAS sample in our analysis.

We functionally annotated the GWS loci in all four SUTs to explore plausible underlying biological processes. We found that associations for AUD were enriched in genes (including *CNOT4*) involved in regulating synapse structure and activity, an enrichment not previously observed in the AUD GWAS ^6^. In addition, we find evidence for neuronal differentiation in smoking initiation, in line with findings in the original SMK GWAS ^7^. In the PPI analysis, we observed significantly enriched protein interaction networks for MTAG-AUD and MTAG-SMK. In addition, we observed multiple interactions between novel genes identified by MTAG and previously identified SUT-associated genes, which provide biological support for the MTAG results.

In PRS analyses, the PRS_MTAG_ outperformed the PRS_GWAS_ for all SUTs. This was of particular importance for the CUD PRS, where the PRS_MTAG_ was significantly associated with the diagnosis, whereas the PRS_GWAS_ was not. As PRS for many traits are being considered as potential biomarkers for disorders ^46^, the use of MTAG may yield more powerful PRS without having to recruit larger samples for GWAS. However, careful evaluation is required to assess the broader impact of MTAG-based PRS on phenotypic associations beyond the primary phenotype. A loss of specificity may result from the inclusion of multiple genetically correlated traits, potentially confounding PheWAS of MTAG-PRS.

In summary, in an MTAG analysis of four SUTs we identified 19 novel loci and, in an independent dataset, found that the associations of PRS_MTAG_ with relevant traits were more significant than with PRS_GWAS_. As the size of GWAS samples continues to increase, MTAG analyses could provide a complementary method that leverages more powerful GWAS to boost the findings of risk variants for genetically correlated traits in which case ascertainment is more challenging, thereby enhancing our understanding of the biology underlying these phenotypes.

## Data Availability

Summary statistics used in this work are publicly available.

